# Health Economic Analysis of the Integrated Cognitive Assessment Tool to Aid Dementia Diagnosis in the United Kingdom

**DOI:** 10.1101/2022.05.09.22274765

**Authors:** Judith Shore, Chris Kalafatis, Mohammad Hadi Modarres, Seyed-Mahdi Khaligh-Razavi

## Abstract

**Objectives:** The aim of this study was to develop a comprehensive economic evaluation of the Integrated Cognitive Assessment (ICA) tool compared with standard cognitive tests when used for dementia screening in primary care and for initial patient triage in memory clinics.

**Methods:** ICA was compared with standard of care comprising a mixture of cognitive assessment tools over a lifetime horizon and employing the UK health and social care perspective. The model combined a decision tree to capture the initial outcomes of the cognitive testing with a Markov structure that estimated long-term outcomes of people with dementia. Quality of life outcomes were quantified using quality-adjusted life years (QALYs). Both costs and QALYs were discounted at 3.5% per annum and cost-effectiveness was assessed using a threshold of £20,000 per QALY gained.

**Results:** ICA dominated standard cognitive assessment tools in both the primary care and memory clinic settings. Introduction of the ICA tool was estimated to result in a lifetime cost saving of approximately £147 and £283 per person in primary care and memory clinics, respectively. QALY gains associated with early diagnosis were modest (0.0019 in primary care and 0.0035 in memory clinic). The net monetary benefit of ICA introduction was estimated at £184 in the primary care and £368 in the memory clinic settings.

**Conclusions:** Introduction of ICA as a tool to screen primary care patients for dementia and perform initial triage in memory clinics could be cost saving to the UK public health and social care payer.

## Introduction

Dementia is defined as an acquired loss of cognition that affects everyday function.^1^ It is an umbrella term for a number of specific medical conditions, including Alzheimer’s disease (AD),^2^ which is perhaps the most studied subtype of dementia. In 2019, the global number of individuals who lived with dementia was estimated at 57.4 million and, largely due to population growth and ageing, this figure is expected to approximately triple by 2050 to reach 152.8 million.^3^

In the UK, 885,000 people were estimated to live with dementia in 2019; the majority of them (84.7%) residing in England.^4^ The number of people with dementia in the UK has been projected to increase to 1.6 million by 2040, including 1.35 million people in England alone.^4^ The economic burden of dementia in the country is substantial, with the total cost of care estimated at £34.7 billion across the UK in 2019, of which publicly or privately funded social care constituted 45% (£15.7 billion), informal care 40% (£13.9 billion), and health care 14% (£4.9 billion).^4^ By 2040, the total costs of dementia care have been projected to rise 2.7-fold from the 2019 estimates, to reach approximately £94.1 billion.^4^

Currently there are no disease-modifying therapies are available in the UK and the existing pharmacologic interventions work symptomatically.^5, 6^ In addition, non-pharmacological interventions such as cognitive stimulation, cognitive rehabilitation, and occupational therapy are recommended to promote independence and well-being in people with dementia.^6^ However, there are still potential benefits to diagnosing dementia early. Firstly, there is some evidence beginning to emerge that early treatment for AD delays cognitive decline.^7^ Secondly, early awareness of diagnosis facilitates informed decisions, e.g. related to financial and legal planning or future care needs.^8^ Finally, with the advent of disease-modifying therapies (DMTs), which target the pathophysiology of AD to delay its progression^9^, the importance of early dementia diagnosis will be growing in years to come and early diagnosis may provide the opportunity to participate in research.

Early diagnosis of dementia at a stage where the cognitive impairment is still mild can be achieved in the primary care setting using an established cognitive assessment instrument.^8, 10^ In the UK, individuals with suspected dementia are subsequently referred to a memory clinic to establish the subtype of dementia based on further diagnostic testing and initiate therapy as appropriate.^10^ Cognitive testing is often also employed during initial triage in the memory clinic.^11^ The COVID-19 pandemic, however, necessitated conducting cognitive assessment remotely, despite unclear validity and reliability of such assessments using currently available tools.^12^

The Integrated Cognitive Assessment (ICA), trademarked as CognICA(tm), is a brief, language independent, self-administered, computerized cognitive test, which uses an explainable artificial intelligence model to improve its accuracy of cognitive impairment diagnosis.^13-15^ The tool aims to facilitate and streamline dementia diagnosis in the National Health Service (NHS) and was assessed in the real-world Accelerating Dementia Pathway Technologies (ADePT) study.^11^ The aim of the current analysis was to develop a comprehensive economic evaluation of the ICA tool compared with standard cognitive tests used for dementia screening in the primary care setting and for initial patient triage in the memory clinic setting.

## Methods

In the base case analysis, the ICA tool was compared to standard cognitive testing in the primary care setting, comprising the Mini-Mental State Examination (MMSE), General Practitioner Assessment of Cognition (GPCOG), the 6-Item Cognitive Impairment Test (6CIT), the Abbreviated Mental Test score (AMTS), the Montreal Cognitive Assessment (MoCA) and a small proportion received a mix of other tests. A scenario analysis was also conducted in people with suspected dementia that have been referred to a memory clinic, where the ICA tool was tested against standard care comprising the Addenbrooke’s Cognitive Examination-III (ACE-III) and the Montreal Cognitive Assessment (MoCA). Quality of life outcomes were quantified using quality-adjusted life years (QALYs). Both costs and QALYs were discounted at 3.5% per annum in line with National Institute for Health and Care Excellence (NICE) guidance.^16^ The NICE reimbursement threshold of £20,000 per QALY gained was used to assess the cost-effectiveness of the ICA tool over a lifetime horizon.

### Model structure

Model structure is summarized in Figure 1. The structure was informed by the Sussex partnership memory clinic care pathway and a targeted literature review. The model employed a decision tree to capture the initial outcomes of the cognitive testing, followed by a Markov structure to capture long-term outcomes after the initial test. A 1-year cycle length was used to capture dementia progression in the Markov model. The perspective of the analysis was that of the UK NHS and Personal Social Services (PSS).

**Figure 1.**
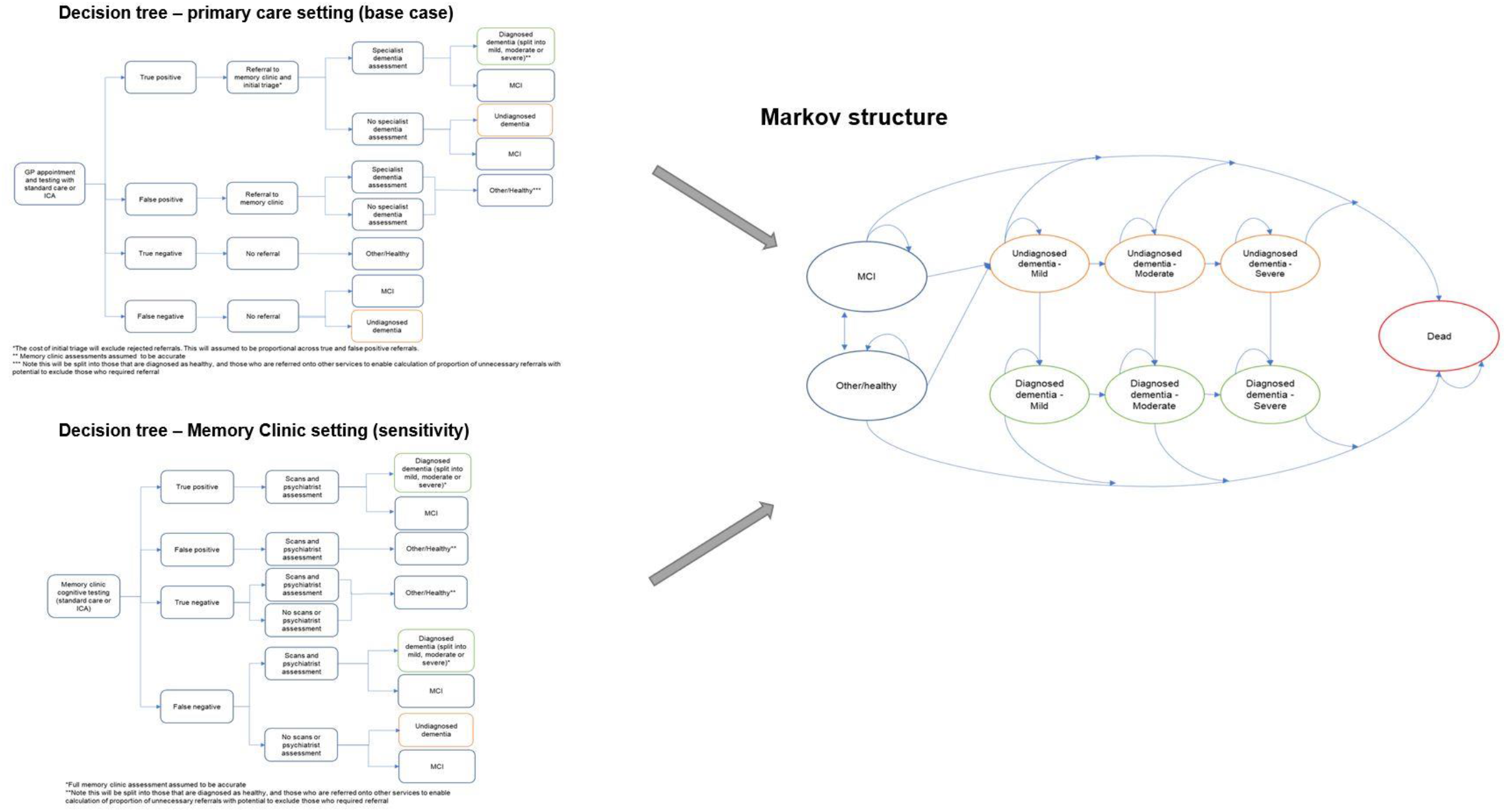
Structure of the model including the two alternative decision tree components (left) and the Markov component (right).

### Clinical and quality of life inputs

A summary of model inputs is provided in Table 1.

**Table 1.** Model inputs applied in the base case and scenario analyses.

| Parameter | Input | Source |
| --- | --- | --- |
| Base case (Primary care setting) |  |  |
| Epidemiology |  |  |
| Proportion of patients attending GP with symptoms | 0.6% | 35 |
| Proportion of patients refusing assessment in a GP clinic | 6.28% | Figures from 2018 to 2019 used so as to minimize impact of the COVID-19 pandemic |
| Prevalence of dementia in those attending the GP | 6.4% | 36 |
| Prevalence of MCI in those attending the GP | 5.5% | 37 |
| Testing outcomes |  |  |
| ICA tool |  |  |
| Sensitivity for MCI | 79% | ADePT study<sup>11</sup>. Data on file |
| Specificity for MCI | 79% |  |
| Sensitivity for Dementia | 97% |  |
| Specificity for Dementia | 79% |  |
| Standard care cognitive tests |  |  |
| Mini-mental state examination (MMSE) |  |  |
| Sensitivity for MCI | 51% | 18 |
| Specificity for MCI | 75% |  |
| Sensitivity for Dementia | 59% |  |
| Specificity for Dementia | 85% |  |
| Proportion of patients receiving test | 26% |  |
| General Practitioner assessment of cognition (GPCOG) |  |  |
| Sensitivity for MCI | 52% | 18 |
| Specificity for MCI | 82% |  |
| Sensitivity for Dementia | 60% |  |
| Specificity for Dementia | 93% |  |
| Proportion of patients receiving test | 21% |  |
| 6-Item Cognitive Impairment Test (6CIT) |  |  |
| Sensitivity for MCI | 66% | 18 |
| Specificity for MCI | 70% |  |
| Sensitivity for Dementia | 88% |  |
| Specificity for Dementia | 78% |  |
| Proportion of patients receiving test | 29% |  |
| Abbreviated mental test score (AMTS) |  |  |
| Sensitivity for MCI | 66% | Assumption |
| Specificity for MCI | 70% |  |
| Sensitivity for Dementia | 81% | 38 |
| Specificity for Dementia | 84% |  |
| Proportion of patients receiving test | 7% | 18 |
| Montreal Cognitive Assessment (MoCA) |  |  |
| Sensitivity for MCI | 80% | 39 |
| Specificity for MCI | 81% |  |
| Sensitivity for Dementia | 91% | 40 |
| Specificity for Dementia | 81% |  |
| Proportion of patients receiving test | 6% | 18 |
| Other |  |  |
| Sensitivity for MCI | 63% |  |
| Specificity for MCI | 76% | Average of other tests in use |
| Sensitivity for Dementia | 79% |  |
| Specificity for Dementia | 85% |  |
| Proportion of patients receiving test | 11% | 18 |
| Weighted average of standard care tests used in model |  |  |
| Sensitivity for MCI | 60% | Calculated using information above |
| Specificity for MCI | 75% |  |
| Sensitivity for Dementia | 73% |  |
| Specificity for Dementia | 84% |  |
| Transition probabilities and mortality |  |  |
| Relative risk of death for undiagnosed/diagnosed dementia compared with general population | 1.82 | 18, 19 |
| Relative risk of death for MCI compared with general population | 1.00 | 18 |
| Underlying dementia severity at point of testing – Proportion mild | 78% | 17 |
| Underlying dementia severity at point of testing – Proportion moderate | 16% |  |
| Underlying dementia severity at point of testing – Proportion severe | 6% |  |
| Proportion of patients with mild undiagnosed dementia being diagnosed each year | 9% | Sensitivity of standard care tests for each severity band combined with proportion going for test<sup>18, 41</sup> |
| Proportion of patients with moderate undiagnosed dementia being diagnosed each year | 13% |  |
| Proportion of patients with severe undiagnosed dementia being diagnosed each year | 86% |  |
|  |  | 90% of severe patients were assumed to go for testing |
| Progression of dementia from mild to moderate each year – undiagnosed | 25.9% | 17<br>Mild to moderate patients grouped as moderate |
| Progression of dementia from mild to moderate each year – diagnosed | 16.0% |  |
| Progression of dementia from moderate to severe each year – undiagnosed | 18.7% |  |
| Progression of dementia from moderate to severe each year – diagnosed | 11.6% |  |
| Yearly transition from MCI to mild undiagnosed dementia | 4.9% | 18 |
| Yearly transition from MCI to 'other/healthy' | 16.0% |  |
| Yearly transitions from 'other/healthy' to mild undiagnosed dementia | Age up to 69 years: 0.7%<br>Age 70 to 74: 1.1%<br>Age 75 to 79: 1.4%<br>Age 80 to 84: 2.2%<br>Age 85+: 6.3% |  |
| Yearly transitions from 'other/healthy' to MCI | Age up to 69 years: 1.2% |  |
|  | Age 70 to 74:<br>1.8%<br>Age 75 to 79:<br>3.3%<br>Age 80 to 84:<br>3.2%<br>Age 85+: 2.3% |  |
| Quality of life |  |  |
| Utility for 'healthy/other' health state | 0.749 to 0.645 | Population norm values by age<sup>20</sup><br>used with<br>assumption of linear<br>change between age<br>groups |
| Utility decrement for MCI | -0.06 | 42 |
| Utility decrements for diagnosed dementia |  |  |
| Mild | -0.125 | 43 |
| Moderate | -0.235 | 43 |
| Severe | -0.305 | 44 |
| Utility decrements for undiagnosed dementia |  |  |
| Mild | -0.129 | 30 |
| Moderate | -0.242 |  |
| Severe | -0.314 |  |
| Cost of testing |  |  |
| Cost of GP time (per minute) | £3.60 | 45 |
| Cost of practice nurse (per minute) | £0.62 | 45 |
| Total cost of laboratory dementia screening tests per patient | £8.31 | 45 |
| GP time taken to undertake/interpret ICA tool test (minutes) | 2 | Data on file |
| GP time taken to undertake/interpret standard care tests – weighted average (minutes) | 8 | 18, 46-49 |
| Practice nurse time to undertake ICA tool test (minutes) | 10 | Data on file |
| Practice nurse time to undertake standard care test – weighted average (minutes) | 0 | 18, 46-49 |
| ICA tool cost per test | £10* | Data on file |
| Standard care cost per test – weighted average | £0.26 | 18<br>AMTS, MoCA and ‘Other’ were assumed to be free of charge. |
| Total cost of ICA tool | £31.67 | Calculated using above inputs |
| Total cost of standard care tests | £36.72 |  |
| Health state costs |  |  |
| Diagnosed dementia |  |  |
| Mild | £9,699 | 31<br>Method used based on Getsios et al.<sup>21</sup><br>Costs of treatment based on NICE guidelines and costed using BNF.<sup>10, 24</sup> |
| Moderate | £32,418 |  |
| Severe | £34,301 |  |
| Undiagnosed dementia |  |  |
| Mild | £8,973 | 31 |
| Moderate | £31,692 |  |
| Severe | £34,055 |  |
| Mild cognitive impairment (ICA tool) | £0.00 | 32 |
| Mild cognitive impairment (standard care) | £0.00 |  |
| Other/healthy | £0.00 |  |
| **Scenario (Memory clinic setting)** |  |  |
| Epidemiology |  |  |
| Proportion of patients attending the GP with symptoms | 0.6% | 35 |
| Referral rate to memory clinic | 9.8% |  |
| Proportion of patients declining referral to memory clinic | 6.7% |  |
| Prevalence of dementia in those attending the memory clinic | 63% | 50 |
| Prevalence of MCI in those attending the memory clinic | 18% |  |
| **Testing outcomes** |  |  |
| ICA tool |  |  |
| Sensitivity for MCI | 79% | ADePT study<sup>11</sup>, Data on file |
| Specificity for MCI | 79% |  |
| Sensitivity for Dementia | 97% |  |
| Specificity for Dementia | 79% |  |
| Standard care cognitive tests |  |  |
| The Addenbrooke's Cognitive Examination – III (ACE-III) |  |  |
| Sensitivity for MCI | 75% | 51 |
| Specificity for MCI | 89% |  |
| Sensitivity for Dementia | 94% |  |
| Specificity for Dementia | 83% |  |
| Proportion of patients receiving test | 50% | Assumption |
| Montreal Cognitive Assessment (MoCA) |  |  |
| Sensitivity for MCI | 80% | 39 |
| Specificity for MCI | 81% |  |
| Sensitivity for Dementia | 91% | 40 |
| Specificity for Dementia | 81% |  |
| Proportion of patients receiving test | 50% | Assumption |
| Weighted average of standard care tests used in model |  |  |
| Sensitivity for MCI | 78% | Calculated using<br>information above |
| Specificity for MCI | 85% |  |
| Sensitivity for Dementia | 93% |  |
| Specificity for Dementia | 82% |  |
| Cost of testing |  |  |
| Cost of nurse time per minute (weighted average of bands 5 to 7) | £1.13 | Assumed 80% at band 5, 10% at band 6 and 10% at band 7<sup>45</sup> |
| Nurse time taken to undertake ICA test (minutes) | 10 | Data on file |
| Nurse time taken to undertake standard care cognitive tests (minutes) – weighted average | 12.5 | 49 |
| ICA tool cost per test | £10* | Data on file |
| Total cost of ICA testing | £21.27 | Calculated using<br>above inputs |
| Total cost of standard care cognitive testing | £14.08 |  |
| Cost of further tests and scans in a memory clinic | £961 | 45 |
| Abbreviations: GP: general practitioner; ICA: Integrated cognitive assessment; MCI: mild cognitive impairment |  |  |

Sensitivity and specificity of the ICA tool for diagnosing mild cognitive impairment (MCI) and dementia were derived from the ADePT study.^11^ The preliminary results from this study informed the performance of the ICA tool in comparison with specialist diagnosis obtained at the memory clinic, i.e. the accuracy of ICA for referral of patients to a memory clinic. Final results of the ADePT study, including the performance of ICA for triage of patients in the memory clinic before entering subsequent diagnostic pathway, are expected in the second quarter of 2022.

Underlying dementia severity in year 1 (i.e., at the point of testing) was based on the Teipel et al. model assessing the cost-effectiveness of donepezil for AD^17^ and was assumed to be the same regardless of diagnosis status (i.e. in people with dementia who are diagnosed vs undiagnosed). The proportions of the modelled population that were diagnosed with mild, moderate, and severe dementia in subsequent years were taken from the literature and assumed to be equal between the two testing arms. Progression of undiagnosed and diagnosed dementia was derived from Teipel et al.^17^ Transition probabilities from MCI to mild undiagnosed dementia and between the ‘other/healthy’ health state and MCI were sourced from a study by Tong et al. assessing the cost-effectiveness of different cognitive screening tests for use in the primary care setting.^18^

Mortality was modelled using UK general population data for individuals in the ‘healthy/other’ health state and relative risks were applied for people with dementia to capture increased risk of death. The relative risk of death with dementia was assumed to be the same regardless of whether the condition was diagnosed or undiagnosed, in line with a previous study.^19^ People with MCI were assumed to die at the same rate as the general population, based on the study by Tong et al.^18^

Utility values were estimated using UK population norms^20^ with literature-derived decrements applied to dementia health states.

### Cost and resource use inputs

The model was populated using epidemiological and cost and resource use data from official NHS and PSS data sources, wherever available (cost year 2019). Costs of testing included in the model comprised staff time to conduct the cognitive tests, laboratory testing, and the costs of further assessment and scans at a memory clinic, in addition to the cost of the ICA.

The cost of ICA comprised a one-off implementation fee of £2,000, followed by a minimum monthly fee of £1,000 for 100 tests with each test thereafter charged at £10. Implementation fees were charged per trust and annuitised in the model. Sourcing of iPads or tablets, which are required for ICA to operate, were not included assuming these would be already available at the primary care practice or memory clinic settings. Staff time for training on the use if the ICA tool was also not included in the model.

Based on the methods used by Getsios et al.,^21^ the costs of undiagnosed and diagnosed dementia were assumed to be the same, except for the cost of treatment which was applied only to those who had been diagnosed. This assumption can be considered conservative, since potential cost savings associated with earlier diagnosis (e.g., due to delay in admission to care homes) have been reported in the literature.^22, 23^ Treatment costs were based on NICE guidelines and costed using the BNF.^10, 24^

### Model outputs

The following outcomes were assessed for each testing arm of the model and compared between arms: total costs per person, total QALYs per person, total number of diagnoses per modelled population, total number of unnecessary referrals per modelled population, and total number of scans/assessments at a memory clinic per modelled population. Head-to-head incremental comparison between the ICA tool and standard care was based on the incremental cost-effectiveness ratio (ICER), the incremental net health benefit (NHB); and the incremental net monetary benefit (NMB).

### Sensitivity analysis

In order to account for uncertainty around the input parameter values, one-way deterministic sensitivity analysis (DSA) was included in the model. The ranges used to vary the parameters were based on confidence intervals, or assumptions were made where these were not available. A probabilistic sensitivity analysis (PSA) was also implemented, whereby, depending on the parameter of interest, Dirichlet, gamma, lognormal or beta distributions were fitted to uncertain parameters to generate the input values for each iteration. The standard errors used to generate probabilistic values were derived from reported 95% confidence intervals wherever possible.

Where this information was not available (e.g., for some costs), the standard error was assumed to be equal to 25% of the mean value. The appropriate number of iterations for PSA was determined by observing the number of iterations required for the average results to stabilize.

### Scenario analysis

At present, data on the sensitivity and specificity of ICA used for initial triage in the memory clinic are limited and, consequently, this application of the tool was assessed in a scenario analysis assuming equivalent sensitivity and specificity as in the base case analysis. Model inputs specific to this scenario are listed in Table 1 below the base case inputs.

Two additional scenarios are presented in the Supplementary Material: a scenario considering the use of the ICA tool for remote initial assessment of patients with symptoms of dementia and a scenario considering the use of the ICA tool for remote monitoring of MCI patients.

## Results

### Base case model results (primary care setting)

The ICA tool dominated standard care comprising currently used cognitive tests, with an estimated NMB of £184 per person (Table 2). The use of the ICA tool in the primary care setting to facilitate preliminary diagnosis of dementia and referral to memory clinics resulted in cost savings of around £147 per person over a lifetime time horizon. This equated to a cost saving to the NHS of approximately £44,222,760 when considering the total population in a given year who may benefit from the introduction of the ICA tool. A small incremental QALY benefit of 0.0019 per person was also estimated as a result of earlier dementia diagnosis.

**Table 2.** Summary of base case results (primary care setting)

|  | ICA tool | Standard care | Incremental |
| --- | --- | --- | --- |
| Costs |  |  |  |
| **Total costs** | **£7,784,633,971** | **£7,828,856,731** | **-£44,222,760** |
| Cost per patient | £25,879 | £26,026 | -£147 |
| QALYs |  |  |  |
| **Total QALYs** | **1,673,814** | **1,673,252** | **562.24** |
| QALYs per patient | 5.56 | 5.56 | 0.0019 |
| Other outcomes |  |  |  |
| ICER | Dominant |  |  |
| NMB | £184 |  |  |
| NHB | 0.01 |  |  |
| **Total number of referrals** | **131,365** | **121,020** | **10,345** |
| **Total number of diagnoses** | **16,661** | **12,584** | **4,078** |
| Number of unnecessary referrals | 93,302 | 90,883 | 2,419 |
| Abbreviations: ICA: integrated cognitive assessment; ICER: incremental cost effectiveness ratio;<br>NHB: net health benefit; NMB: net monetary benefit; QALYs: quality-adjusted life-years |  |  |  |

Although there were additional costs associated with introducing the tool (implementation costs and test fees for ICA and increased costs to memory clinics for increased referrals), these costs were outweighed by the reduction in staff costs associated with conducting the initial cognitive tests and a reduction in dementia care costs associated with earlier diagnosis and therefore delayed progression to more severe dementia states (Table 3).

**Table 3.** Detailed cost breakdown in the primary care setting

|  | **ICA tool** | **Standard care** | **Incremental** |
| --- | --- | --- | --- |
| Implementation costs | £57,778 | £0 | £57,778 |
| Cost of initial testing | £9,528,113 | £11,045,037 | -£1,516,925 |
| Cost of referral and memory clinic triage | £1,850,055 | £1,704,366 | £145,689 |
| Cost of further assessments at memory clinic | £56,151,268 | £48,745,905 | £7,405,362 |
| Other/healthy | £0 | £0 | £0 |
| MCI | £0 | £0 | £0 |
| Undiagnosed dementia mild | £1,133,307,831 | £1,188,128,893 | -£54,821,062 |
| Undiagnosed dementia moderate | £1,757,257,606 | £1,940,889,563 | -£183,631,957 |
| Undiagnosed dementia severe | £310,758,991 | £355,334,234 | -£44,575,243 |
| Diagnosed dementia mild | £662,607,427 | £579,875,947 | £82,731,480 |
| Diagnosed dementia moderate | £2,015,535,625 | £1,832,211,993 | £183,323,632 |
| Diagnosed dementia severe | £1,837,579,278 | £1,870,920,793 | -£33,341,514 |
| **Total** | **£7,784,633,971** | **£7,828,856,731** | **-£44,222,760** |
| Abbreviations: ICA: Integrated cognitive assessment; MCI: mild cognitive impairment |  |  |  |

### Sensitivity analysis

Detailed results of the sensitivity analysis are presented in the Supplemental Material (Supplementary Figure 1). Key cost-effectiveness drivers included the progression of undiagnosed dementia (transition from undiagnosed mild to undiagnosed moderate dementia), and the relative risk of death for people with undiagnosed dementia vs the general population. Each of these parameters could change the direction of the results when varied alone. PSA results indicated that the probability of the ICA tool being cost-effective was 72% at the £20,000 per QALY threshold.

### Scenario analysis (Memory clinic setting)

When introduced as a triage tool in memory clinic services, ICA dominated standard care and NMB was estimated at £368 per person. Employing the ICA tool saved approximately £298 per person over a lifetime time horizon, which equated to an overall cost saving to the NHS of around £8,702,393 (Table 4). Similar to the primary care setting, there was a small QALY gain of 0.0035 per person associated with earlier diagnosis of dementia.

**Table 4.** Summary of scenario analysis results (memory clinic setting)

|  | ICA tool | Standard care | Incremental |
| --- | --- | --- | --- |
| Costs |  |  |  |
| **Total costs** | **£3,037,436,676** | **£3,046,139,068** | **-£8,702,393** |
| Cost per patient | £103,929 | £104,227 | -£298 |
| QALYs |  |  |  |
| **Total QALYs** | **121,722** | **121,620** | **101.92** |
| QALYs per patient | 4.16 | 4.16 | 0.0035 |
| Other outcomes |  |  |  |
| ICER | Dominant |  |  |
| NMB | £368 |  |  |
| NHB | 0.02 |  |  |
| **Total number of referrals** | **24,103** | **22,799** | **1,304** |
| **Total number of diagnoses** | **17,860** | **17,031** | **829** |
| Number of unnecessary referrals | 1,955 | 1,572 | 383 |

The increase in initial costs due to the increased number of individuals being tested for dementia was outweighed by savings due to diagnosing dementia earlier and therefore delaying progression to more severe states of the disease (Table 5). The costs in the memory clinic setting were much higher compared with the primary care setting, due to substantially higher prevalence of dementia in this setting.

**Table 5.** Detailed cost breakdown in the memory clinic setting

|  | **ICA tool** | **Standard care** | **Incremental** |
| --- | --- | --- | --- |
| Implementation costs | £57,778 | £0 | £57,778 |
| Cost of triage | £621,540 | £411,600 | £209,940 |
| Cost of further testing | £23,163,267 | £21,910,146 | £1,253,122 |
| Other/healthy | £0 | £0 | £0 |
| MCI | £0 | £0 | £0 |
| Undiagnosed dementia mild | £56,989,956 | £68,128,825 | -£11,138,869 |
| Undiagnosed dementia moderate | £102,458,394 | £139,769,825 | -£37,311,432 |
| Undiagnosed dementia severe | £19,741,864 | £28,798,927 | -£9,057,063 |
| Diagnosed dementia mild | £468,696,059 | £451,886,186 | £16,809,873 |
| Diagnosed dementia moderate | £1,369,566,366 | £1,332,317,582 | £37,248,784 |
| Diagnosed dementia severe | £996,141,452 | £1,002,915,979 | -£6,774,527 |
| **Total** | **£3,037,436,676** | **£3,046,139,068** | **-£8,702,393** |

DSA results revealed that the most influential parameter was the sensitivity of the ICA tool for diagnosing dementia in the memory clinic setting, which was varied by an arbitrary range of ±20% pending availability of the relevant data from the ADePT study. This parameter, alongside the two parameters already described as influential in the base case analysis (transition probability from undiagnosed mild dementia to undiagnosed moderate dementia and the relative risk of death for people with undiagnosed dementia vs the general population) could alter the direction of the results when each of them was varied alone. In terms of PSA results, the probability of the ICA tool being cost-effective in the memory clinic setting was 63% at the £20,000 per QALY threshold.

## Discussion

The ICA tool dominated cognitive assessment tests currently used in both the primary care and memory clinic settings. Introduction of the ICA tool was estimated to result in a lifetime cost saving to the NHS of approximately £147 per person undergoing cognitive assessment in the primary care setting and £283 per person being triaged in the memory clinic setting. Although the model predicted an increase in costs from increased referrals and specialist assessments, these were outweighed by a reduction in the staff costs associated with initial cognitive testing and a reduction in the costs of treatment and care for people with dementia achieved through earlier diagnosis. Increasing referrals and improving diagnosis rates is in line with the Department of Health national dementia strategy^25^.

The potential for earlier diagnosis of dementia through the use of the ICA tool is particularly important given that the first DMT for AD was approved last year by the Food and Drug Administration (FDA)^9^ and the AD pipeline appears rich with other DMT candidates.^26^ Potential introduction of cost-effective DMTs for AD and other types of dementia would further improve the case for early diagnosis of the condition and therefore increase the cost-effectiveness of more accurate screening^27, 28^ using tools such as the ICA. The progression of dementia for those who have mild undiagnosed dementia in relation to those who have been diagnosed (i.e. are assumed to be receiving treatment) is a key driver in the model and could change the direction of the results if there is no difference in progression of dementia with treatment. Therefore, further research in this area will also improve the robustness of the model results.

The results from the present study align reasonably well with previous UK-based analyses of similar decision problems published in recent years. Tong et al. reported on a cost-effectiveness analysis undertaken to assess the use of different cognitive screening tests for diagnosing dementia and MCI in primary care.^18^ The results of Tong et al. employing the health and social care perspective were similar to this analysis, ranging from a cost of £6.93 to a saving of £185 per person over a lifetime horizon, depending on the cognitive test used.^18^ However, the incremental QALYs reported were more optimistic than in the present analysis,^18^ potentially due to the differences in sources informing utility data. Getsios et al conducted an economic evaluation comparing early assessment and treatment for AD with early assessment and no treatment, as well as treatment without early assessment.^21^ Although the estimated cost savings from early assessment and treatment were much higher than that in this analysis (£2,135 per person when compared to treatment without early assessment and £3,593 when compared to no early assessment and no treatment),^21^ some important differences in model design and assumptions between the Getsios et al. model and the present study could explain these differences. The Getsios et al. analysis estimated the potential savings from assessing and treating all individuals with dementia early and assumed 100% assessment accuracy, whereas this study estimated cost savings resulting from only a proportion of individuals benefiting from earlier diagnosis and treatment i.e., those diagnosed with the ICA tool that would have not been diagnosed with standard cognitive tests. Therefore, the cost savings estimated in this analysis could be expected to be more modest.

Although the present model concerns a highly relevant issue considering the ageing UK population and the potential advent of DMTs for dementia, it should be noted that the analysis was subject to several limitations. Firstly, the ADePT study which informed inputs related to ICA accuracy did not include a comparison between ICA and other cognitive assessment instruments, although it did compare the outputs from the ICA tool with further testing undertaken in a memory clinic which can be considered the gold standard. Since the sensitivity and specificity inputs for standard cognitive tests were based on information extracted from the literature with no attempts to validate them, the current study should be considered a naïve comparison. Additionally, the ADePT study did not include participants who were not referred by their GPs to memory clinics, so its population is somewhat different than the population included in the model.

There is a paucity of data around the influence of diagnosis on the health care costs and quality of life associated with dementia. Therefore, the health care costs associated with diagnosed and undiagnosed dementia for each severity level were assumed to be the same, except for treatment costs assigned only to individuals who had received a dementia diagnosis. The potential benefits associated with earlier diagnosis of dementia have been described in the literature^8^, however, no studies generalizable to the UK NHS that quantify these benefits in terms of the relative difference in health care costs between individuals with and without a diagnosis could be identified. Similarly, the quality of life benefits associated with diagnosis do not seem to be widely reported across prior cost-effectiveness analyses and literature reviews.^21-23, 29^ The base case utility value employed in the present study was based on Gomes et al., who reported EQ-5D data at baseline and 6 months following diagnosis.^30^ However, 6 months after diagnosis can be considered a short time frame in a chronic condition, so employing this value would lead the model to understate quality of life benefits if quality of life gains were made past the first 6 months following diagnosis. The uncertainty associated with the cost and quality of life impact of dementia diagnosis could affect the cost-effectiveness of the ICA tool, as there were fewer individuals with undiagnosed dementia in the ICA arm of the model, so that increasing the cost or disutility applied to undiagnosed dementia would improve the cost-effectiveness of the ICA tool, while reducing these costs or utility decrements would decrease the cost-effectiveness of the tool.

The model also did not account for all potential costs. The cost of supplying iPads or tablets, which would be required for the ICA tool to be used in practice, was not included, assuming that these would be available at GP practices and/or memory clinics. In reality, most clinics and practices can be expected to have access to such devices, so that the costs of any additional purchases would likely be negligible. In addition, no costs were assigned to individuals in the MCI health states who were assumed to receive no regular monitoring or treatment in line with current treatment pathways. If such costs were included, they would be higher in the ICA tool arm of the model, so any benefits associated with monitoring would need to outweigh these additional costs.

The model employed the NHS and PSS perspective; consequently, the costs associated with informal care and other out-of-pocket costs were not included, even though much of the cost and burden of care falls on the person with dementia and their family.^31^ Anderson et al. estimated that the costs of unpaid care and other costs falling on individuals vary between £9,711 for individuals with severe dementia to £17,917 for those with milder dementia.^32^ This equates to anywhere between approximately 25% of the total costs estimated per year for individuals with moderate and severe dementia up to 70% for those with milder dementia who may receive less formal care.^32^ However, there is substantial uncertainty associated with costing informal care. The aforementioned analysis by Tong et al. estimated only a small change in results when informal care costs were included.^18^ It should also be noted that, in addition to informal care costs, the costs of lost productivity (for people with dementia or their caregivers) were also not considered, even though by 2040 these costs are estimated to reach £1.3 billion to businesses in England.^33^ If informal care costs and productivity costs were to be included in this analysis this would further improve the cost-effectiveness of the ICA tool.

Quality of life of caregivers or family members was also not considered, despite the substantial impact that caring for people with dementia has on them.^34^ Including these costs and quality of life benefits would likely improve the case for the adoption of the ICA tool because higher costs associated with dementia, particularly more severe states, could be expected to result in higher cost savings (provided the costs for diagnosed individuals are not substantially higher than those for undiagnosed individuals). It should be noted, however, that the cost-effectiveness threshold used in this analysis would no longer be relevant when considering the societal perspective because it is intended to represent the opportunity cost of funding an intervention using NHS and PSS budgets. Therefore, interpretation of the cost-effectiveness results is more difficult when considering this wider perspective.

A further limitation of the model is that it does not consider the wider impact of introducing the ICA tool on health care system capacity. Introducing a cognitive test that has increased sensitivity but decreased specificity compared with standard care would lead to increased number of referrals to memory clinics, thus having an impact on the clinic capacity. Similarly, introducing a test with increased sensitivity but reduced specificity in memory clinics as a triage tool would likely lead to more patients being routed to further testing. The costs of expanding the capacity of these services or any other services that may be impacted by an increase in diagnosed cases of dementia has not been considered in this analysis, nor has the impact of the likely increase in waiting times to access the services.

Finally, this model only considers the use of the ICA tool in place of current cognitive tests used in clinical practice. The ICA tool may have potential other uses including remote testing or monitoring of patients, although further clinical evidence and economic analyses would be needed to evaluate this.

## Conclusion

Introduction of the ICA tool within the NHS as a diagnostic tool for cognitive impairment and dementia in place of cognitive assessment tools currently used in primary care could be cost saving to the NHS. The potential cost savings were even greater when the tool was evaluated as a triage tool in memory clinics; future studies are needed to collect more real-world data in memory clinic settings to investigate the generalization of the results across wider populations in such settings.

## Data Availability

All data produced in the present work are contained in the manuscript

## Acknowledgments

Medical writing and editorial assistance with the preparation of this manuscript was provided by Karolina Badora, PhD. Development of the model was overseen by Michelle Green, MSc. Assistance with designing for the ADePT study protocol was provided by Dr Naji Tabet and Panos Apostolou.

