## Supplementary Material for "Health Economic Analysis of the Integrated Cognitive Assessment Tool to Aid Dementia Diagnosis in the United Kingdom"

### Results of deterministic sensitivity analyses

The tornado diagram listing the top 15 most influential input parameters in the primary care setting (base case) analysis is presented in Supplementary Figure 1. The corresponding results for the memory clinic setting (scenario) are presented in Supplementary Figure 2. Parameters related to progression of dementia and relative risk of death in people with dementia were the most influential in the primary care setting; however, in the memory clinic setting, sensitivity of the ICA tool for dementia had the most influence on model results.

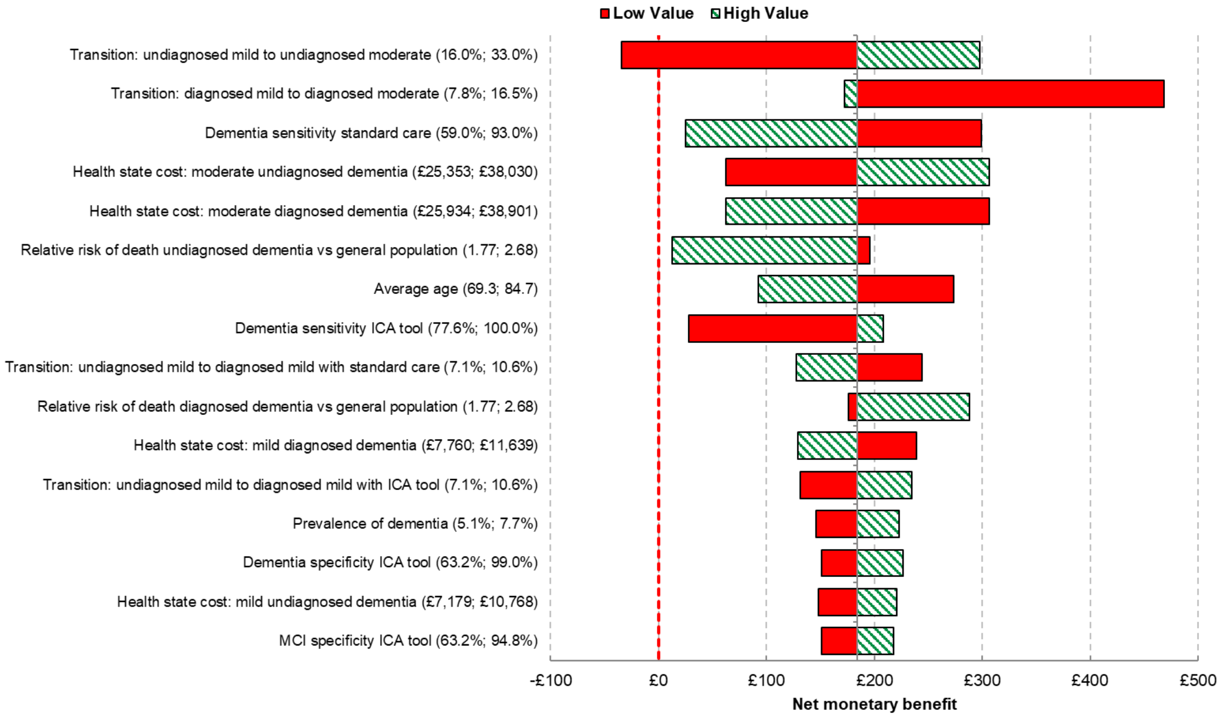


Supplementary Figure 1. Tornado diagram for the primary care setting (base case) analysis

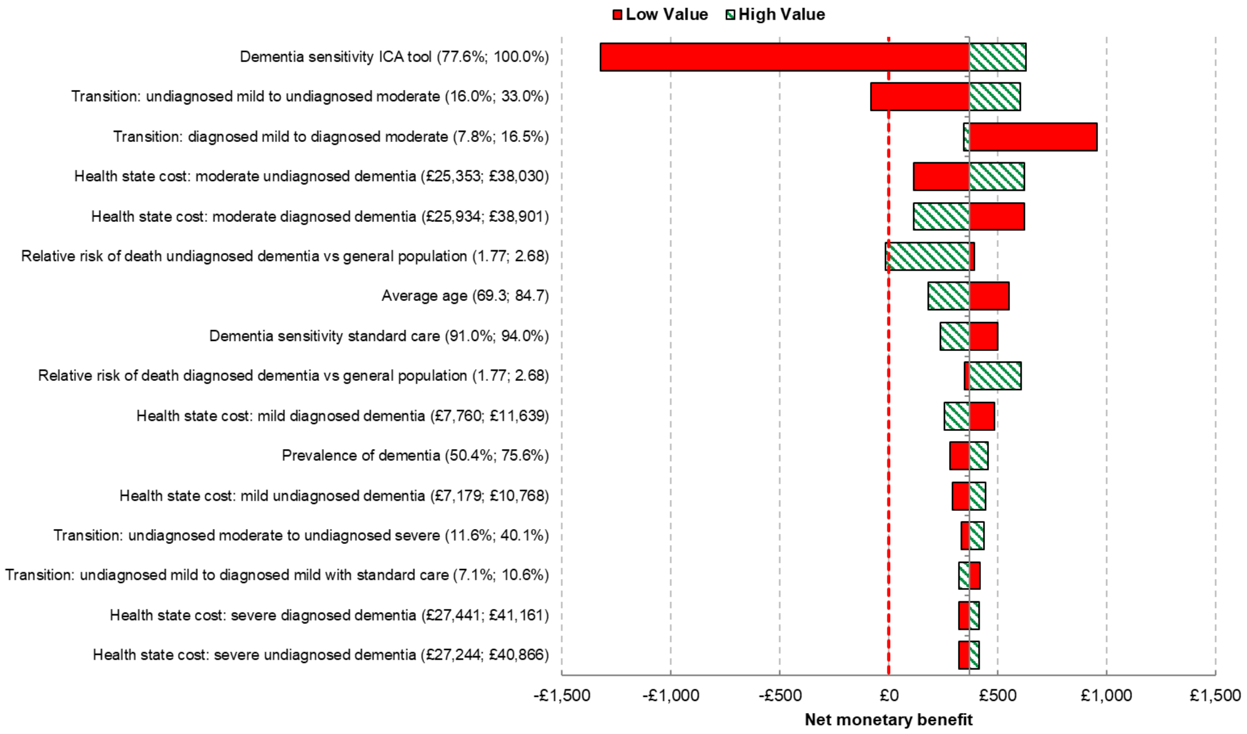


Supplementary Figure 2. Tornado diagram for the memory clinic setting (scenario) analysis

### Additional scenario analyses

#### *Methods of the additional analyses*

Additional scenarios assessed 1) the use of the ICA tool for remote initial assessment of patients with symptoms of dementia and 2) the use of the ICA tool for remote monitoring of MCI patients. Both scenarios applied to the primary care setting.

The ICA tool could be potentially used for remote cognitive assessment of patients in the primary care setting. While relevant evidence has not yet been generated, the nature of the tool makes it suitable for this application. Remote testing of primary care patients could free up resources by reducing staff time associated with testing for dementia.  In this scenario, patients were tested remotely with the ICA tool and, as such, the cost of the nurse time for administering the test was removed. This resulted in the total cost of the ICA test being reduced to £25.51 (vs

£31.67 in the base case). It should be noted, however, that no additional time was included for contacting the patient to discuss results and therefore this scenario can be considered optimistic.

Another potential use of the ICA tool could be the remote monitoring of primary care patients diagnosed with MCI, to facilitate early detection of progression to dementia. Again, no evidence in this area has been generated to date, so the scenario should be considered exploratory. In this scenario, patients in the MCI health state were not split into diagnosed and undiagnosed, and therefore the costs of monitoring in the model were also applied to those patients who remained undiagnosed. The following costs were included for the MCI health state for the ICA tool arm only: a cost of £25 to reflect an annual remote test using the ICA tool, and a cost of £50 to reflect the costs of memory clinic assessments for patients referred for further testing each year (based on the costs of memory clinic triage and further testing and the proportion of patients with MCI expected to progress to dementia each year). In addition, to reflect the accelerated diagnosis of patients who progress from MCI to mild undiagnosed dementia, the transition probability from mild undiagnosed dementia to mild diagnosed dementia with the ICA tool was amended to assume that approximately 75% (vs approximately 22% in the base case) of patients were being tested each year. It should be noted that no other changes to the clinical pathway that may be necessary to administer remote testing for MCI patients were considered within this scenario analysis. Similarly, the costs associated with any increases in testing in memory clinics were not fully considered, and neither were administrative costs associated with following up on patients to ensure they had undertaken the testing as required. Therefore, the results of the scenario should be considered optimistic.

***Additional scenario results***

As shown in Supplementary Table 1, conducting cognitive testing of primary care patients remotely using the ICA tool could further increase the cost savings seen with the ICA tool in the base case. However, the costs of follow up calls or appointments to discuss testing results with the patient were not included in this scenario, so this represents a very optimistic estimate.

Supplementary Table 1. Results of the remote cognitive testing scenario

| | **ICA tool** | **Standard care** | **Incremental** |
|---|---|---|---|
| Costs | | | |
| **Total costs** | **£7,782,778,952** | **£7,828,856,731** | **-£46,077,780** |
| Cost per patient | £25,872 | £26,026 | -£153 |
| QALYs | | | |
| **Total QALYs** | **1,673,814** | **1,673,252** | **562** |
| QALYs per patient | 5.56 | 5.56 | 0.0019 |
| Other outcomes | | | |
| ICER | Dominant | | |
| NMB | £191 | | |
| NHB | 0.01 | | |
| Abbreviations: ICA, Integrated Cognitive Assessment; ICER, incremental cost-effectiveness ratio; NHB, net health benefit; NMB, net monetary benefit; QALY, quality-adjusted life year | | | |

Remotely monitoring patients with MCI in primary care with the ICA tool also showed the potential to increase cost savings estimated from use of the tool in the base case (Supplementary Table 2). It should be noted that this scenario is also optimistic, and the true costs of monitoring MCI patients may be higher. In this scenario, only the costs of testing were considered, and administrative costs associated with setting up such a monitoring system were not included.

Supplementary Table 2. Results of the scenario assessing remote monitoring of patients with MCI

| | **ICA tool** | **Standard care** | **Incremental** |
|---|---|---|---|
| Costs | | | |

| | | | |
|---|---|---|---|
| **Total costs** | **£7,677,135,879** | **£7,828,856,731** | **-£151,720,852** |
| Cost per patient | £25,521 | £26,026 | -£504 |
| QALYs | | | |
| **Total QALYs** | **1,674,974** | **1,673,252** | **1,722** |
| QALYs per patient | 5.57 | 5.56 | 0.0057 |
| Other outcomes | | | |
| ICER | Dominant | | |
| NMB | £619 | | |
| NHB | 0.03 | | |
| Abbreviations: ICA, Integrated Cognitive Assessment; ICER, incremental cost-effectiveness ratio; NHB, net health benefit; NMB, net monetary benefit; QALY, quality-adjusted life year | | | |
